# Lower risk of dementia with Lewy bodies among users of glycolysis enhancing drugs in a new user, active comparator design

**DOI:** 10.1101/2023.12.05.23299455

**Authors:** Alexander Hart, Georgina Aldridge, Qiang Zhang, Nandakumar S. Narayanan, Jacob E. Simmering

## Abstract

**Background:** Terazosin, doxazosin, and alfuzosin (Tz/Dz/Az) are α-1 adrenergic receptor antagonists that also bind to and activate a key ATP-producing enzyme in glycolysis. Recent work has suggested a potentially neuroprotective effect of use of Tz/Dz/Az in Parkinson’s disease in both animal and human studies. We investigated neuroprotective effects of Tz/Dz/Az in a closely related disease, dementia with Lewy bodies (DLB).

**Methods:** We used a new user active comparator design in the merative Marketscan database to identify men with no history of DLB who were newly started on Tz/Dz/Az or two comparator medications. Our comparator medications were other drugs commonly used to treat benign prostatic hyperplasia that do not increase ATP: the α-1 adrenergic receptor antagonist tamsulosin or 5α-reductase inhibitors (5ARI). We matched the cohorts on propensity scores and duration of follow-up. We followed the matched cohorts forward to estimate the hazard of developing DLB.

**Results:** Men who were newly started on Tz/Dz/Az had lower hazard of developing DLB then matched men taking tamsulosin (HR=0.60, p<0.001) or 5ARI (HR=0.73, p=0.012) while the hazard in men taking tamsulosin was similar to that of men taking 5ARI (HR=1.17, p=0.12). These results were robust to several sensitivity analyses.

**Conclusions:** These data demonstrate that Tz/Dz/Az in decreases the risk of DLB. When combined with the literature of Tz/Dz/Az on PD, our findings suggest that glycolysis-enhancing drugs may be broadly protective in neurodegenerative synucleinopathies. These observational associations must be further evaluated for causality in future randomized trials.

## Introduction

Dementia with Lewy Bodies (DLB) is a progressive neurodegenerative disorder characterized by cognitive impairment and parkinsonism. Additional symptoms such as dysautonomia, sleep disorders, hallucinations, and cognitive fluctuations may also occur. (1) The incidence of DLB has been estimated to be 0.5-1.6 per 1000 people per year and as DLB is implicated in 3.2-7.1% of dementia cases. (2) As age is a primary risk factor, DLB incidence will increase as the population ages. (2) Current pharmacotherapy for DLB focuses on mitigation of symptoms, largely use the use of acetylcholine esterase inhibitors and levodopa. To date, there are no known preventative or disease-modifying treatments for DLB. Finding modifiable risk factors and preventive treatments has the potential to reduce the morbidity and mortality of this disease. (3)

Recently, impaired energetics has been explored as a potential target for prevention and treatment of neurodegenerative diseases, especially Parkinson’s disease (PD). The α-1 blockers terazosin (Tz), doxazosin (Dz), and alfuzosin (Az; collectively Tz/Dz/Az) were found to have an additional target: phosphoglycerate kinase 1 (PGK1), the first ATP producing enzyme in glycolysis. Tz/Dz/Az bind PGK1 and Tz has been shown to increase ATP in cells, animal models, and in humans. (4) In several preclinical PD models (5) and pharmacoepidemiologic cohort studies in humans, (6-9) Tz successfully reduced the rate of neuronal loss and disease morbidity. Pre-clinical studies have suggested a more generalized protection against neurodegeneration, including in amyotrophic lateral sclerosis (10) and Alzheimer’s disease. (11) Given these results and considering the pathological similarities between DLB and PD, we asked if glycolysis-enhancing drugs offered neuroprotection in DLB.

Tz/Dz/Az are widely used in older men, chiefly to manage symptoms related to benign prostatic hyperplasia (BPH). (12) DLB is a disease of aging and disproportionality affects men (13) yielding an alignment between the people at highest risk of DLB and the primary users of these medications. Importantly, tamsulosin, another α-1 blocker, has similar clinical indications and effectiveness (14) but, crucially, does not bind to PGK1 and does not increase ATP. These features make tamsulosin is an ideal active comparator – the observational approximation of a placebo – as men taking tamsulosin or Tz/Dz/Az are likely similar on many observed and unobserved characteristics but tamsulosin is not expected to have any effect on PGK1. For additional rigor, we investigated an additional comparator also used to treat BPH: the 5α reductase inhibitors (5ARI) finasteride and dutasteride. These drugs have a distinct mechanism from α-1 blockers and have no known increase in neuronal ATP. (15)

Using these two comparators (tamsulosin and 5ARI), we sought to estimate the association between use of Tz/Dz/Az and the later development of DLB using a retrospective, observational cohort study of health insurance claims.

## Methods

### Data Source

The Merative Marketscan Commercial Claims and Encounters (CCAE) and Medicare Supplemental and Coordination of Benefits (MDCR) databases from 2001 to 2017 were utilized for this study. Using the health insurance claims, we identified men with a dispensing claim for Tz/Dz/Az, tamsulosin, finasteride (5ARI) or dutasteride (5ARI). We utilized both outpatient and inpatient clinical information on all diagnoses submitted on bills to insurance companies that were present in the above databases. The existing database was previously deidentified and thus not considered human subjects research and is exempt from IRB review. This database and our general analytical framework has previously been used to study the hazard of developing PD and PD-related impairment. (5, 7, 16)

### Cohort Construction

We defined our cohort using a new user active comparator design, the standard for pharmacoepidemiologic studies. (17, 18) Specifically, we identified individuals newly started on medications of interest (Tz/Dz/Az) or active comparators (tamsulosin, 5ARI). We identified dispensing events by matching the National Drug Code numbers included for those medications in the 2015 Redbook. For each individual, we identified the first observed dispensing event of the BPH medications and excluded any enrollee who took more than one medication group. To ensure the “new user” element of our design, we required at least 365 days of enrollment with prescription drug coverage before the first observed dispensing date. Additionally, we required at least one day of follow-up after the medication dispensing date.

To reduce the potential for selection effects due to discontinuation, we additionally required at least a second dispensing claim in the in the first year after the index date. Our reasoning was that individuals who only have a single dispensing event are unlikely to be actually taking the medication while those who have a refill are likely users.

As these drugs are primarily used for BPH we excluded female enrollees to reduce heterogeneity in the study cohort as female individuals cannot develop BPH. Both BPH and DLB are exceedingly rare under the age of 40 so we required enrollees to be at least 40 years old at the first observed dispensing date. As our interest is on the changes in risk of developing DLB associated with use of these medications, we excluded anyone who had a diagnosis of DLB on or before the medication start date.

### DLB Case Definition

Patients were defined as having DLB if a claim with one or more DLB diagnosis code (ICD-9-CM: 3318.2 or ICD-10CM: G31.83) in any setting (inpatient or outpatient) was made. A diagnosis of PD with an associated dementia/cognitive impairment code without a DLB diagnosis was not counted as a case. We defined our outcome date as the first date that the person is diagnosed with DLB in any setting (inpatient, outpatient, emergency department).

### Propensity Score Matching

To reduce differences between the study cohorts, we used propensity score matching. The propensity score included the year of medication start; age at medication start; incidence rate of outpatient visits, mean number of unique diagnoses recorded per outpatient visit, incidence rate of unique outpatient diagnoses during the lookback period; the incidence of hospitalization during lookback; a diagnosis of BPH (ICD-9-CM: 600.xx or ICD-10-CM: N40.x) on or before the index date; whether prostate specific antigen (PSA) levels were measured (CPT: 84152, 84153, 84154) or were diagnosed as abnormal (ICD-9-CM: 790.93, ICD-10-CM: R97.2, R97.20, R97.21); a diagnosis of slow urinary stream (ICD-9-CM: 788.62, ICD-10-CM: R39.12); whether a uroflow study was done (ICD-9-CM procedure code: 89.24, ICD-10-CM procedure code: 4A1D75Z, CPT: 51736, 51741); whether a cystometrogram was collected (ICD-9-CM procedure code: 89.22, ICD-10-CM procedure code: 4A0D7BZ, 4A0D8BZ, 4A1D7BZ, 4A1D8BZ, CPT: 51725, 51726); a diagnosis of orthostatic hypotension (ICD-9-CM: 458.0 or ICD-10-CM: I95.1); a diagnosis of other hypotension (ICD-9-CM: 458.1, 458.2x, 458.8, 458.9 or ICD-10-CM: I95.0, I95.2, I95.3); the 30 Elixhauser comorbidity flags, as revised by AHRQ. (19)

For the year of medication start, we included a series of dummy variables as we expected changes in medication use patterns, the rate of diagnosis of DLB, and the sample included in the Marketscan database over time. Insurance companies enter and leave the Marketscan project creating potentially large changes in the Marketscan-enrollee-universe at the end/start of a year. Including dummy variables for year accounts for these problems. We used splines of all the continuous variables (e.g., rate of outpatient encounters during lookback, age) to allow for non-linear responses and estimated the propensity score using generalized additive models with a logistic link.

We used a two-step matching algorithm. First, we required the time from the medication start date to the end of enrollment to be similar (+/- 90 days) to ensure balance in time-at-risk between the cases and controls. Second, within the set of possible matches with similar follow-up, we used greedy nearest-neighbor matching based on the estimated log odds.(20) To ensure matches were high quality, we imposed a caliper equal to 20% of the pooled standard deviation of the log odds. We matched 1:1 without replacement. In the event of multiple equally good matches, we selected the matching control observation at random. We then had three matched cohorts for the three comparisons of interest: TZ/DZ/AZ versus tamsulosin, TZ/DZ/AZ versus 5ARI, and tamsulosin versus 5ARI.

### Assessing Propensity Score Match Balance

Groups were compared before and after matching on all variables included in the propensity score model. We utilized Cohen’s d to assess balance. Cohen’s d is a common standardized measure of effect size between groups. We predefined the absolute value of Cohen’s d of <0.1 as indicating minimal difference between the various covariates.(21, 22)

### Analysis

We estimated the survival function non-parametrically using the Kaplan Meier estimator and tested for equality of the survival curves using the log-rank test. We quantified the difference in survival using the semi-parametric Cox proportional hazards regression using robust standard errors clustered by the pairing generated through the propensity score matching. The proportional hazards assumption was assessed using the Schoenfeld residuals.

### Sensitivity Analyses

First, we estimated Cox proportional hazards models with time-interacted covariates to mitigate potential violations of the proportional hazards assumption. A major concern would be if the first year or two had very different hazard ratios than the later years, suggesting treatment selection was endogenous to the future outcome. This may happen if people were sorted between the treatments by their (unobserved) risk of orthostatic hypotension, which is a prognostic factor for future DLB development. If this were the case, we may see a strong association during DLB and treatment choice initially (first 1-2 years) but no association with longer time differences.

Second, we restricted our sample to only men with a diagnosis of BPH, elevated PSA, a history of PSA measurement or other diagnosis or procedure suggestive of urinary dysfunction. Relative to the overall cohort, this subsample should have less heterogeneity in treatment indication, albeit at significant sample size costs. Our estimates should be consistent between the propensity score matched sample and this narrow subsample if the groups are truly balanced.

Third, we included the requirement that men have 2 or more claims for dispensing of the medication in order to count as users of the medication. Men with a single claim are likely not users as these medications are generally used for long time durations as opposed to acute or one-off treatments (e.g., antibiotics). However, this poses a threat: men who discontinue due to side effects on the first court are excluded under this rule. If discontinuation due to side effects varies by medication and is correlated with our outcome of DLB, as might be the case with some side effects like orthostatic hypotension, then this rule may introduce a bias in favor of Tz/Dz/Az that is spurious. We repeat our matching and analysis using an intent-to-treat structure including anyone who has ever had a dispensing event for the medication.

Fourth, we varied the start of follow-up from immediately upon starting the mediation (the index date) to 1, 2, or 3 years after the start of the index date. If the hazard ratio is substantially different – if it starts low and goes to 1 – this is suggestive that treatment selection is endogenous to future DLB risk and the observed results are due to selection effects.

All analyses were performed in R 4.2.2 (23), the icd package was used to calculate the Elixhauser comorbidities, the propensity score generalized additive models were estimated using the mgcv package (24, 25), and survival analyses were performed using the survival (26) package. The R code for data manipulation and analysis are available at https://github.com/iacobus42/dlb-tz-paper.

## Results

We analyzed 121,358 pairs of men in the Tz/Dz/Az versus tamsulosin analysis after matching, 65,436 pairs of men in the Tz/Dz/Az versus 5ARI analysis after matching, and 79,798 pairs of men in the tamsulosin versus 5ARI analysis after matching. All groups were well balanced (the absolute value of Cohen’s d or Cohen’s w less than 0.10) on all included covariates with good overlap in the estimated scores (Tables S1 to S3). Summaries of the duration of follow-up and cumulative incidence are reported for each of the three cohorts in Table 1.

**Table 1:** Number of people, duration of follow-up, number of cases, and cumulative incidence by cohort, medication before and after propensity score matching.

|  | Medication | Before Matching |  |  |  | After Matching |  |  |  |
| --- | --- | --- | --- | --- | --- | --- | --- | --- | --- |
|  |  | No. of People | Person-Years | No. of Cases | Cases Per 10k Per Year | No. of People | Person-Years | No. of Cases | Cases Per 10k Per Year |
| Tz/Dz/Az versus Tamsulosin | Tz/Dz/Az | 126,313 | 373,989 | 195 | 5.21 | 121,358 | 363,311 | 193 | 5.28 |
|  | Tamsulosin | 437,045 | 1,195,288 | 1,286 | 10.76 | 121,358 | 364,945 | 323 | 8.85 |
| Tz/Dz/Az versus 5ARI | Tz/Dz/Az | 126,313 | 373,989 | 195 | 5.21 | 65,436 | 199,561 | 111 | 5.56 |
|  | 5ARI | 80,158 | 248,090 | 193 | 7.78 | 65,436 | 199,755 | 152 | 7.61 |
| Tamsulosin versus 5ARI | Tamsulosin | 437,045 | 1,195,288 | 1,286 | 10.76 | 79,798 | 246,086 | 224 | 9.1 |
|  | 5ARI | 80,158 | 248,090 | 193 | 7.78 | 79,798 | 246,194 | 192 | 7.8 |

Kaplan Meier survival curves for the raw and matched cohorts are shown in Figure 1. Prior to adjustment, we found statistically significant lower hazards of DLB in men who were taking Tz/Dz/Az versus tamsulosin (HR = 0.65, p < 0.001) or 5ARI (HR = 0.73, p < 0.001). Critically, the hazards in men taking tamsulosin were not significantly different than men taking 5ARI, implying that tamsulosin was reasonable null effect comparison group (HR = 1.12, p = 0.10). This pattern persisted after matching with men taking Tz/Dz/Az having consistently lower hazards of DLB than men taking tamsulosin (HR = 0.60, p < 0.001) or 5ARI (HR = 0.73, p = 0.01) while men taking tamsulosin had similar hazards to men taking 5ARI (HR = 1.17, p = 0.12). Full details are reported in Table 2.

**Table 2:** Estimated hazard ratio and 95% confidence intervals by cohort before and after propensity score matching.

|  |  | Unadjusted |  |  | Propensity Score Matched |  |  |
| --- | --- | --- | --- | --- | --- | --- | --- |
| Treatment | Reference | HR | 95% CI | p-value | HR | 95% CI | p-value |
| Tz/Dz/Az | Tamsulosin | 0.48 | 0.41,<br>0.56 | < 0.001 | 0.60 | 0.50,<br>0.71 | < 0.001 |
| Tz/Dz/Az | 5ARI | 0.67 | 0.55,<br>0.82 | < 0.001 | 0.73 | 0.57,<br>0.93 | 0.012 |
| Tamsulosin | 5ARI | 1.40 | 1.20,<br>1.62 | < 0.001 | 1.17 | 0.96,<br>1.42 | 0.12 |

**Figure 1:**
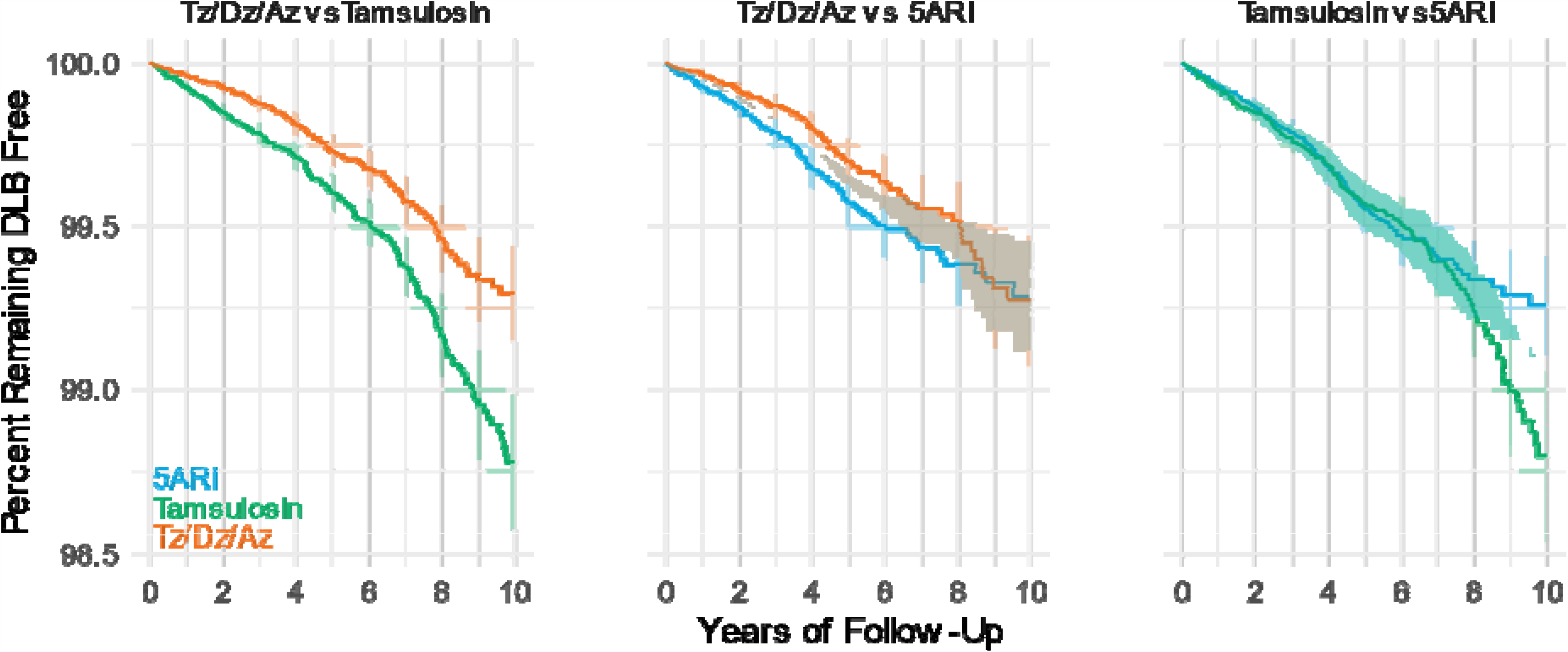
Kaplan-Meier survival curves for Tz/Dz/Az versus tamsulosin, Tz/Dz/Az versus 5ARI, and tamsulosin versus 5ARI. All comparisons are after propensity score matching. Shaded regions denote 95% CIs.

Inspection of the Schoenfeld residuals indicated no meaningful correlation with time for Tz/Dz/Az versus tamsulosin (r = -0.25; 95% CI: -0.10, 0.08; p = 0.803) but potential violations of the proportional hazards assumption Tz/Dz/Az versus 5ARI (r = 0.14; 95% CI: 0.02, 0.26; p = 0.022) and tamsulosin versus 5ARI (r = 0.13; 95% CI: 0.03, 0.22; p = 0.009). We repeated our analysis using time-interactions to quantify this concern. The overall estimate and the time-interaction estimate are very similar for at least 5 years of follow-up; after 5-years, the comparisons including 5ARI change with the overall estimate understating the time-interacted HR, Table S4. However, these later time periods appear to have poorly estimated hazard ratios, likely due to relatively sparse data at extremely long follow-up periods.

Restricting the analysis to only those men with a BPH diagnosis, PSA measurement, or LUTS-related procedural code yielded directionally consistent results. Our primary comparison – men taking Tz/Dz/Az versus tamsulosin – still indicated statistically significantly lower hazard in men who were taking Tz/Dz/Az (HR = 0.73, 9% CI: 0.55-0.97). Men taking Tz/Dz/Az had non-significantly lower hazard of DLB then men taking 5ARI (HR = 0.87, 95% CI: 0.63-1.20) while men on tamsulosin had similar hazard as men taking 5ARI (HR = 1.23, 95% CI: 0.95-1.59). In each of these analyses, there is approximately a 25% decrease in sample size when requiring a diagnosis of BPH, measurement of PSA, or other urinary-symptom-related procedure.

Intent-to-treat analysis including all men who ever had a dispensing event for one of the study drugs had estimated results almost identical to the main analysis. Men taking Tz/Dz/Az had lower hazards of DLB then men taking tamsulosin (HR = 0.69; 95% CI: 0.59, 0.81; p < 0.001) or 5ARI (HR = 0.75; 95% CI: 0.61, 0.93; p = 0.010) while the hazard was similar between men taking tamsulosin compared to men taking 5ARI (HR = 1.13; 0.94, 1.35; p = 0.19).

Finally, varying the start of follow-up resulted in directionally consistent estimated hazard ratios for all three comparisons. Increasing delays of follow-up reduce the sample size leading to broader 95% CIs and reduced power with 50% or more reductions in sample size within 3 years. The sample size, number of cases, and estimated HRs are reported in Table 3. Taken together, these data provide evidence for a neuroprotective role of Tz/Dz/Az in the development of DLB.

**Table 3:** Effect of delayed start of follow-up on estimated hazard ratio.

| Comparison | Delay in Years | No. of Cases | No. of People | No. of Pairs | HR | 95% CI |
| --- | --- | --- | --- | --- | --- | --- |
| Tz/Dz/Az versus Tamsulosin | 0 | 514 | 242,696 | 121,348 | 0.60 | 0.50, 0.71 |
|  | 1 | 386 | 175,968 | 87,984 | 0.67 | 0.55, 0.82 |
|  | 2 | 385 | 121,908 | 60,954 | 0.80 | 0.64, 1.01 |
|  | 3 | 242 | 88,908 | 44,454 | 0.72 | 0.55, 0.92 |
| Tz/Dz/Az versus 5ARI | 0 | 260 | 130,866 | 65,433 | 0.70 | 0.55, 0.90 |
|  | 1 | 200 | 95,754 | 47,877 | 0.84 | 0.63, 1.10 |
|  | 2 | 161 | 68,158 | 34,079 | 0.89 | 0.65, 1.22 |
|  | 3 | 113 | 49,414 | 24,707 | 0.92 | 0.63, 1.32 |
| Tamsulosin versus 5ARI | 0 | 412 | 159,554 | 79,777 | 1.16 | 0.95, 1.41 |
|  | 1 | 328 | 118,624 | 59,312 | 1.33 | 1.07, 1.65 |
|  | 2 | 240 | 84,724 | 42,362 | 1.20 | 0.93, 1.55 |
|  | 3 | 152 | 61,732 | 30,866 | 0.90 | 0.66, 1.23 |

## Discussion

We extend prior research studying showing a potential protective effect of Tz/Dz/Az use in PD to the closely related disease of DLB. The protective association with Tz/Dz/Az use is seen in both comparisons against another alpha-blocker tamsulosin and in the clinically but not pharmacologically related 5ARIs. Also of significance, there was no significant difference between men who took tamsulosin or 5ARI after matching, making it unlikely either of these medications could be having harmful alternative effect, such as causing dementia.

These results are in broad agreement with the emerging literature for a neuroprotective effect in users of Tz/Dz/Az. Our research group has previously reported neuroprotective effects in animal models of Parkinson’s disease (5) and Parkinson’s disease dementia (16) as well as finding negative associations between Tz/Dz/Az use and Parkinson’s disease symptoms (5), development of Parkinson’s disease dementia (16), and lower hazard of developing Parkinson’s disease in both the United States and Denmark. (27) A pilot clinical trial confirmed increased ATP levels in the brain among people with Parkinson’s disease who were given Tz. (28) Our findings of an observational protective association in users of Tz/Dz/Az were replicated by independent groups using health insurance claims in the United States (9) and Canada. (8) Pre-clinical animal models of Alzheimer’s disease in mouse models (11) and amyotrophic lateral sclerosis in zebrafish, mouse, and neuron models. (10) Considering these results along with our findings of a protective association in DLB suggests a potentially broad neuroprotective effect for Tz/Dz/Az. As our hypothesized mechanism is the effect on PGK1 and enhanced metabolism offsetting some of the age-related metabolic impairment, such a broad effect is biologically plausible and expected.

There are functionally no highly effective, affordable, and safe disease-modifying therapy for these fatal neurodegenerative diseases. Emerging monoclonal antibodies for Alzheimer’s disease are likely not cost effective at current prices (29) but there will be considerable pressure for Medicare to cover these medications. Repurposing an existing, FDA approved medication, like Tz/Dz/Az, or targeting PGK1 activity may offer greater effectiveness at lower cost.

Any observational study must address many threats of confounding. Unobserved factors may relate both to the outcome of interest and the choice of medication. Our design, a new user, active comparator design paired with propensity score matching, reduces many threats of confounding and is the gold standard design in pharmacoepidemiology. (17, 18, 30, 31) By using an active comparator, we have a situation where cases or controls could have been started on either the study medication or the control medication. This reduces the risk of unobserved confounders and helps ensure the groups are as homogenous as possible, even before using propensity score matching.

Additionally, using a second, independent control group of men who are newly started on 5ARI allows us to assess the validity of our tamsulosin control. While not as clinically interchangeable with Tz/Dz/Az or tamsulosin, 5ARI are used to primarily used to manage BPH symptoms. By considering men using 5ARIs, we are making a comparison to men with BPH who have opted into pharmacological treatment of their condition. These men likely have more in common with men starting Tz/Dz/Az or tamsulosin than men who either do not have BPH or who do have BPH but elect to opt out of treatment for their symptoms. We found no statistically significant increase in risk in men taking tamsulosin compared to 5ARI suggesting that either tamsulosin is a viable, null effect control medication or that both tamsulosin and 5ARI, despite being very different classes of medication, have a similar effect on DLB by chance. The former interpretation seems more plausible than the latter. This makes the results noted in prior studies finding an increased risk of developing dementia and other synucleinopathies amongst patients who take tamsulosin less likely. (9, 32)

The similarity of the results using time-interacted covariates during the first 5 years or when using a delayed start to follow-up suggest that selection on unobserved factors (e.g., known but not documented orthostatic hypotension) is not driving our result. The estimated effects are present even with a three-year delay. Additionally, our results are nearly identical when using an intent-to-treat analysis and the 2+ dispensing analysis suggesting discontinuations during the first course are not causing selection bias that determines our result. Reducing our sample to only men with a diagnosis of BPH, history of PSA measurement, or LUTS-related procedures yields directionally consistent results, although the sample size reduction of nearly one-third reduces our power in this analysis.

Like all studies ours is not without limitations. One persistent challenge is the relationship between prodromal DLB and lower urinary tract symptoms (LUTS). As autonomic dysfunction is a possible early symptom of DLB, it may be possible that men who will develop DLB in the near future will exhibit urinary retention today. By using an active comparator design we are selecting men who are electing to undergo pharmacological treatment of LUTS symptoms, likely but not exclusively caused by BPH. This mitigates against unobserved differences that may be related to future DLB risk and that would confound our estimates.

Tamsulosin, sharing a mechanism and of similar clinical effectiveness, is a better control medication as a result. The 5ARI are suitable – they are also used to treat BPH – but the longer duration until clinically significant relief (typically 90 days versus immediate for the alpha blockers) suggest they may not be exactly clinically interchangeable. Another important limitation is the poor recording of orthostatic hypotension in the database, which may be a prognostic sign of DLB and is also a reason to prefer tamsulosin or 5ARI over Tz/Dz/Dz. While our intent-to-treat analysis suggests experienced side effects and discontinuation did not bias our result, selection by the provider based on orthostatic risk when selecting treatment may remain present. Lastly there are limitations inherent to the style of research such as lack of randomization, limited ability to assess cause and effect, and limitations to variables present in the data set. We have addressed these challenges through propensity score matching and our sensitivity analyses to uncover any potential bias. Finally, all claims-based analyses are limited by the sensitivity and specificity of diagnostic codes included on billing data. Given the diagnosis of DLB is very exact, it is likely our case definition has very high specificity but potentially low sensitivity. Missing diagnoses of DLB will likely bias our analysis towards the null hypothesis as cases are miscategorized as healthy individuals.

Our study provides supports that enhancing glycolysis with Tz/Dz/Az may reduce the incidence of DLB, a result of extreme importance given the extensive morbidity and mortality of DLB and existing lack of disease modifying therapies. This result aligns with the preclinical and observational support for other neurodegenerative diseases and suggests a broader potential rule of impaired metabolism in neurodegenerative disease. Given the observational nature of this study, we cannot show causality and only estimate associations; future preclinical, prospective observational, and, ultimately, randomized trials are needed to evaluate this finding for causality.

## Data Availability

The merative Marketscan database is used under license from merative that prohibits redistribution of the data. Interested users may contact merative to obtain a license for their use.

https://github.com/iacobus42/dlb-tz-paper

