## Supplemental Materials for "Lower risk of dementia with Lewy bodies among users of glycolysis enhancing drugs in a new user, active comparator design"

Table S1: Description of the Tz/Dz/Az versus tamsulosin cohort before and after matching using propensity scores. Values reported are the number and percent (for categorical measures) and the median, lower, and upper quartiles (for continuous measures). The Std. Diff. column reports standardized measures of differences between the Tz/Dz/Az and tamsulosin groups. For continuous measures, the standardized difference is described using Cohen’s d. For categorical measures, Cohen’s w is used. Both Cohen’s d and w have roughly the same interpretation with values closer to 0 being smaller differences. By convention, absolute standardized differences smaller than 0.1 are considered small-to-no difference. Before matching, Tz/Dz/Az users had earlier start dates than tamsulosin users and less lookback time but were similar on all other measures. After matching of the estimated standardized differences were below 0.1 with many being 0.01 or smaller.

|  |  | Entire Cohort | | | Propensity Score Matched Cohort | | |
| --- | --- | --- | --- | --- | --- | --- | --- |
|  |  | Tamsulosin | Tz/Dz/Az | Std. Diff. | Tamsulosin | Tz/Dz/Az | Std. Diff. |
| Sample Size, Number of Cases | Number of Enrollees | 437,045 | 126,313 | --- | 121,358 | 121,358 | --- |
|  | Number of Cases of DLB | 1,286 | 195 | --- | 323 | 193 | --- |
| Age, Timing of Index Date, Outcome | Age at Index Date | 62  (56-71) | 62  (55-70) | 0.06 | 62  (55-70) | 52  (55-70) | 0.01 |
|  | Index Date Year | 2012  (2008-14) | 2009  (2006-12) | 0.48 | 2009  (2006-12) | 2009  (2006-12) | 0.01 |
| Duration of Enrollment Time | Lookback Time (Years) | 3.6  (2.0-6.3) | 2.9  (1.7-5.2) | 0.24 | 3.2  (1.9-5.5) | 2.9  (1.8-5.3) | 0.06 |
|  | Follow-Up Time (Years) | 1.9  (0.9-3.8) | 2.0  (0.9-4.2) | 0.09 | 2.1  (0.9-4.3) | 2.1  (0.9-4.3) | 0.00 |
| Baseline Utilization and Complexity | Rate of Inpatient Events Per Year | 0.00  (0.00-0.22) | 0.00  (0.00-0.13) | 0.08 | 0.00  (0.00-0.14) | 0.00  (0.00-0.14) | 0.01 |
|  | Rate of Outpatient Events Per Year | 10  (5-18) | 9  (5-16) | 0.07 | 9  (5-17) | 9  (5-16) | 0.01 |
|  | Mean Number of Diagnosis Codes Per Outpatient Event | 1.56  (1.27-2.09) | 1.36  (1.18-1.83) | 0.28 | 1.38  (1.19-1.87) | 1.39  (1.19-1.85) | 0.01 |
| Elixhauser Comorbidities on Index Date | Alcohol Abuse | 11,379  (2.6%) | 2,680  (2.1%) | 0.01 | 2,670  (2.2%) | 2,628  (2.2%) | 0.00 |
|  | Anemia | 68,691  (15.7%) | 17,016  (13.5%) | 0.03 | 16,774  (13.8%) | 16,530  (13.6%) | 0.00 |
|  | Blood Loss | 7,232  (1.7%) | 1,514  (1.2%) | 0.02 | 1,512  (1.2%) | 1,492  (1.2%) | 0.00 |
|  | Heart Failure | 42,970  (9.8%) | 10,207  (8.1%) | 0.02 | 10,289  (8.5%) | 9,931  (8.2%) | 0.01 |
|  | Coagulopathy | 18,949  (4.3%) | 3,645  (2.9%) | 0.03 | 3,621  (3.0%) | 3,580  (2.9%) | 0.00 |
|  | Depression | 43.797  (10.0%) | 8,991  (7.1%) | 0.04 | 8,973  (7.4%) | 8,887  (7.3%) | 0.00 |
|  | Diabetes Without Complications | 121,292  (27.8%) | 36,574  (29.0%) | 0.01 | 35,321  (29.1%) | 35,178  (29.0%) | 0.00 |
|  | Diabetes with Complications | 46,695  (10.7%) | 14,387  (11.4%) | 0.01 | 13,721  (11.3%) | 13,825  (11.4%) | 0.00 |
|  | Drug Abuse | 6,690  (1.5%) | 1,331  (1.1%) | 0.02 | 1,322  (1.1%) | 1,315  (1.1%) | 0.00 |
|  | Fluid or Electrolyte Disorders | 56,021  (12.8%) | 13,805  (10.9%) | 0.02 | 13,649  (11.2%) | 13,421  (11.1%) | 0.00 |
|  | HIV | 1,438  (0.3%) | 486  (0.4%) | 0.00 | 489  (0.4%) | 472  (0.4%) | 0.00 |
|  | HTN Without Complications | 271,591  (62.1%) | 87,650  (69.4%) | 0.06 | 84,253  (69.4%) | 83,676  (68.9%) | 0.01 |
|  | HTN With Complications | 57,049  (13.1%) | 20,761  (16.4%) | 0.04 | 20,108  (16.6%) | 19,729  (16.3%) | 0.00 |
|  | Hypothyroidism | 43,432  (9.9%) | 9,809  (7.8%) | 0.03 | 9,665  (8.0%) | 9,610  (7.9%) | 0.00 |
|  | Liver Disease | 24,205  (5.5%) | 4,845  (3.8%) | 0.03 | 4,780  (3.9%) | 4,790  (3.9%) | 0.00 |
|  | Lymphoma | 6,367  (1.5%) | 1,322  (1.0%) | 0.01 | 1,279  (1.1%) | 1,301  (1.1%) | 0.00 |
|  | Metastatic Cancer | 10,716  (2.5%) | 1,555  (1.2%) | 0.03 | 1,620  (1.3%) | 1,547  (1.3%) | 0.00 |
|  | Neurological Disorders | 45,438  (10.4%) | 8,237  (6.5%) | 0.06 | 8,407  (6.9%) | 8,157  (6.7%) | 0.00 |
|  | Obesity | 48,019  (11.0%) | 11,470  (9.1%) | 0.03 | 11,448  (9.4%) | 11,313  (9.3%) | 0.00 |
|  | Paralysis | 10,583  (2.4%) | 2,087  (1.7%) | 0.02 | 2,163  (1.8%) | 2,061  (1.7%) | 0.00 |
|  | Pulmonary Hypertension | 13,310  (3.0%) | 2,458  (1.9%) | 0.03 | 2,590  (2.1%) | 2,433  (2.0%) | 0.00 |
|  | Psychoses | 32,490  (7.4%) | 6,910  (5.5%) | 0.03 | 6,974  (5.7%) | 6,844  (5.6%) | 0.00 |
|  | Peptic Ulcer Disease | 1,709  (0.4%) | 297  (0.2%) | 0.01 | 311  (0.3%) | 295  (0.2%) | 0.00 |
|  | COPD | 104,627  (23.9%) | 24,107  (19.1%) | 0.05 | 23,993  (19.8%) | 23,664  (19.5%) | 0.00 |
|  | Peripheral Vascular Disease | 65,114  (14.9%) | 15,324  (12.1%) | 0.03 | 15,208  (12.5%) | 14,977  (12.3%) | 0.00 |
|  | Renal Disease | 32,280  (7.2%) | 12,096  (9.6%) | 0.04 | 11,642  (9.6%) | 11,460  (9.4%) | 0.00 |
|  | Rheumatoid Arthritis | 22,205  (5.1%) | 4,754  (3.8%) | 0.03 | 4,605  (3.8%) | 4,667  (3.8%) | 0.00 |
|  | Solid Tumor | 73,745  (16.9%) | 13,905  (11.0%) | 0.07 | 13,861  (11.4%) | 13,720  (11.3%) | 0.00 |
|  | Valvular Disease | 62,332  (14.3%) | 14,081  (11.1%) | 0.04 | 14,157  (11.7%) | 13,866  (11.4%) | 0.00 |
|  | Weight Loss | 20,140  (4.6%) | 3,448  (2.7%) | 0.04 | 3,521  (2.9%) | 3,416  (2.8%) | 0.00 |
| Other Comorbidities and Treatment Selection Factors | Orthostatic Hypotension | 5,423  (1.2%) | 944  (0.7%) | 0.02 | 991  (0.8%) | 939  (0.8%) | 0.00 |
|  | Other Hypotension | 13,208  (3.0%) | 2,202  (1.7%) | 0.03 | 2,209  (1.8%) | 2,175  (1.8%) | 0.00 |
|  | PSA Measurement Taken | 249,926  (57.2%) | 72,215  (57.2%) | 0.00 | 69,873  (57.6%) | 69,205  (57.0%) | 0.01 |
|  | Diagnosis of Abnormal PSA | 70,538  (16.1%) | 14,099  (11.2%) | 0.06 | 14,114  (11.6%) | 13,890  (11.4%) | 0.00 |
|  | Diagnosis of Slow Urinary Stream | 13,752  (3.1%) | 2,948  (2.3%) | 0.02 | 3,059  (2.5%) | 2,931  (2.4%) | 0.00 |
|  | Uroflow Study Performed | 28,542  (6.5%) | 7,684  (6.1%) | 0.01 | 7,859  (6.5%) | 7,567  (6.2%) | 0.00 |
|  | Cystometrogram Performed | 3,485  (0.8%) | 937  (0.7%) | 0.00 | 992  (0.8%) | 926  (0.8%) | 0.00 |
|  | Diagnosis of BPH | 183,019  (41.9%) | 42,481  (33.6%) | 0.07 | 42,736  (35.2%) | 41,698  (34.4%) | 0.01 |
|  | Diagnosis of Anxiety | 39,365  (9.0%) | 7,827  (6.2%) | 0.04 | 7,759  (6.4%) | 7,749  (6.4%) | 0.00 |
|  | Diagnosis of Erectile Dysfunction | 45,596  (10.4%) | 11,268  (8.9%) | 0.02 | 11,226  (9.3%) | 11,079  (9.1%) | 0.00 |

Table S2: Description of the Tz/Dz/Az versus 5ARI cohort before and after matching using propensity scores. Values reported are the number and percent (for categorical measures) and the median, lower, and upper quartiles (for continuous measures). The Std. Diff. column reports absolute value of standardized measures of differences between the Tz/Dz/Az and 5ARI groups. For continuous measures, the standardized difference is described using Cohen’s d. For categorical measures, Cohen’s w is used. Both Cohen’s d and w have roughly the same interpretation with values closer to 0 being smaller differences. By convention, standardized differences smaller than 0.1 are considered small-to-no difference. Before matching, Tz/Dz/Az users had earlier start dates than 5ARI users and were more likely to have hypertension while 5ARI were more likely to have abnormal PSA measurements and an explicit diagnosis of BPH. After matching of the estimated standardized differences were below 0.1 with many being 0.01 or smaller.

|  |  | Entire Cohort | | | Propensity Score Matched Cohort | | |
| --- | --- | --- | --- | --- | --- | --- | --- |
|  |  | 5ARI | Tz/Dz/Az | Std. Diff. | 5ARI | Tz/Dz/Az | Std. Diff. |
| Sample Size, Number of Cases | Number of Enrollees | 80,158 | 126,313 | --- | 65,436 | 65,436 | --- |
|  | Number of Cases of DLB | 193 | 195 | --- | 152 | 111 | --- |
| Age, Timing of Index Date, Outcome | Age at Index Date | 62  (54-71) | 62  (55-70) | 0.04 | 62  (54-70) | 62  (54-70) | 0.01 |
|  | Index Date Year | 2010  (2008-13) | 2009  (2006-12) | 0.22 | 2010  (2007-13) | 2010  (2007-13) | 0.03 |
| Duration of Enrollment Time | Lookback Time (Years) | 3.2  (1.9-5.6) | 2.9  (1.7-5.2) | 0.07 | 3.10  (1.8-5.5) | 3.0  (1.8-5.4) | 0.01 |
|  | Follow-Up Time (Years) | 2.2  (1.0-4.4) | 2.0  (0.9-4.2) | 0.05 | 2.2  (1.0-4.4) | 2.2  (0.9-4.4) | 0.00 |
| Baseline Utilization and Complexity | Rate of Inpatient Events Per Year | 0.00  (0.00-0.00) | 0.00  (0.00-0.13) | 0.11 | 0.00  (0.00-0.00) | 0.00  (0.00-0.00) | 0.00 |
|  | Rate of Outpatient Events Per Year | 9  (5-16) | 9  (5-16) | 0.05 | 9  (5-16) | 9  (5-16) | 0.00 |
|  | Mean Number of Diagnosis Codes Per Outpatient Event | 1.40  (1.20-1.87) | 1.36  (1.18-1.83) | 0.04 | 1.40  (1.19-1.87) | 1.39  (1.19-1.85) | 0.02 |
| Elixhauser Comorbidities on Index Date | Alcohol Abuse | 1,152  (1.4%) | 2,680  (2.1%) | 0.02 | 1,051  (1.6%) | 1,079  (1.6%) | 0.00 |
|  | Anemia | 9,308  (11.6%) | 17,016  (13.5%) | 0.03 | 7,762  (11.9%) | 7,674  (11.7%) | 0.00 |
|  | Blood Loss | 913  (1.1%) | 1,514  (1.2%) | 0.00 | 741  (1.1%) | 735  (1.1%) | 0.00 |
|  | Heart Failure | 5,398  (6.7%) | 10,207  (8.1%) | 0.02 | 4,556  (7.0%) | 4,509  (6.9%) | 0.00 |
|  | Coagulopathy | 2,360  (2.9%) | 3,645  (2.9%) | 0.00 | 1,898  (2.9%) | 1,906  (2.9%) | 0.00 |
|  | Depression | 5,527  (6.9%) | 8,991  (7.1%) | 0.00 | 4,642  (7.1%) | 4,687  (7.2%) | 0.00 |
|  | Diabetes Without Complications | 15,970  (19.9%) | 36,574  (29.0%) | 0.10 | 14,164  (21.6%) | 14,190  (21.7%) | 0.00 |
|  | Diabetes with Complications | 5,141  (6.4%) | 14,387  (11.4%) | 0.08 | 4,712  (7.2%) | 4,615  (7.1%) | 0.00 |
|  | Drug Abuse | 576  (0.7%) | 1,331  (1.1%) | 0.02 | 536  (0.8%) | 549  (0.8%) | 0.00 |
|  | Fluid or Electrolyte Disorders | 6,001  (7.5%) | 13,805  (10.9%) | 0.06 | 5,295  (8.1%) | 5,175  (7.9%) | 0.00 |
|  | HIV | 548  (0.7%) | 486  (0.4%) | 0.02 | 373  (0.6%) | 364  (0.6%) | 0.00 |
|  | HTN Without Complications | 42,463  (53.0%) | 87,650  (69.4%) | 0.17 | 36,898  (56.4%) | 36,836  (56.3%) | 0.00 |
|  | HTN With Complications | 7,995  (10.0%) | 20,761  (16.4%) | 0.09 | 7,217  (11.0%) | 7,176  (11.0%) | 0.00 |
|  | Hypothyroidism | 7,062  (8.8%) | 9,809  (7.8%) | 0.02 | 5,620  (8.6%) | 5,529  (8.4%) | 0.00 |
|  | Liver Disease | 2,476  (3.1%) | 4,845  (3.8%) | 0.02 | 2,141  (3.3%) | 2,174  (3.3%) | 0.00 |
|  | Lymphoma | 844  (1.1%) | 1,322  (1.0%) | 0.00 | 692  (1.1%) | 682  (1.0%) | 0.00 |
|  | Metastatic Cancer | 1,108  (1.4%) | 1,555  (1.2%) | 0.01 | 884  (1.4%) | 849  (1.3%) | 0.00 |
|  | Neurological Disorders | 5,252  (6.6%) | 8,237  (6.5%) | 0.00 | 4,342  (6.6%) | 4,276  (6.5%) | 0.00 |
|  | Obesity | 4,530  (5.7%) | 11,470  (9.1%) | 0.06 | 4,184  (6.4%) | 4,078  (6.2%) | 0.00 |
|  | Paralysis | 1,006  (1.3%) | 2,087  (1.7%) | 0.02 | 879  (1.3%) | 899  (1.4%) | 0.00 |
|  | Pulmonary Hypertension | 1,592  (2.0%) | 2,458  (1.9%) | 0.00 | 1,299  (2.0%) | 1,270  (1.9%) | 0.00 |
|  | Psychoses | 4,257  (5.3%) | 6,910  (5.5%) | 0.00 | 3,560  (5.4%) | 3,627  (5.5%) | 0.00 |
|  | Peptic Ulcer Disease | 185  (0.2%) | 297  (0.2%) | 0.00 | 148  (0.2%) | 155  (0.2%) | 0.00 |
|  | COPD | 8,516  (10.6%) | 24,107  (19.1%) | 0.02 | 11,867  (18.1%) | 12,002  (18.3%) | 0.00 |
|  | Peripheral Vascular Disease | 3,543  (4.4%) | 15,324  (12.1%) | 0.02 | 7,133  (10.9%) | 7,033  (10.7%) | 0.00 |
|  | Renal Disease | 2,857  (3.6%) | 12,096  (9.6%) | 0.09 | 3,354  (5.1%) | 3,296  (3.6%) | 0.00 |
|  | Rheumatoid Arthritis | 2,857  (3.6%) | 4,754  (3.8%) | 0.01 | 2,404  (3.7%) | 2,386  (3.6%) | 0.00 |
|  | Solid Tumor | 10,567  (13.2%) | 13,905  (11.0%) | 0.03 | 8,165  (12.5%) | 8,205  (12.5%) | 0.00 |
|  | Valvular Disease | 9,339  (11.7%) | 14,081  (11.1%) | 0.01 | 7,501  (11.5%) | 7,446  (11.4%) | 0.00 |
|  | Weight Loss | 2,322  (2.9%) | 3,448  (2.7%) | 0.00 | 1,892  (2.9%) | 1,875  (2.9%) | 0.00 |
| Other Comorbidities and Treatment Selection Factors | Orthostatic Hypotension | 823  (1.0%) | 944  (0.7%) | 0.01 | 609  (0.9%) | 592  (0.9%) | 0.00 |
|  | Other Hypotension | 1,532  (1.9%) | 2,202  (1.7%) | 0.01 | 1,233  (1.9%) | 1,205  (1.8%) | 0.00 |
|  | PSA Measurement Taken | 48,463  (60.5%) | 72,215  (57.2%) | 0.03 | 38,893  (59.4%) | 38,607  (59.0%) | 0.00 |
|  | Diagnosis of Abnormal PSA | 23,789  (29.7%) | 14,099  (11.2%) | 0.23 | 13,387  (20.5%) | 13,044  (19.9%) | 0.01 |
|  | Diagnosis of Slow Urinary Stream | 1,963  (2.4%) | 2,948  (2.3%) | 0.00 | 1,639  (2.5%) | 1,687  (2.6%) | 0.00 |
|  | Uroflow Study Performed | 5,672  (7.1%) | 7,684  (6.1%) | 0.02 | 4,499  (6.9%) | 4,809  (7.3%) | 0.01 |
|  | Cystometrogram Performed | 546  (0.7%) | 937  (0.7%) | 0.00 | 483  (0.7%) | 530  (0.8%) | 0.00 |
|  | Diagnosis of BPH | 36,166  (45.1%) | 42,481  (33.6%) | 0.12 | 27,327  (41.8%) | 27,707  (42.3%) | 0.01 |
|  | Diagnosis of Anxiety | 5,501  (6.9%) | 7,827  (6.2%) | 0.01 | 4,458  (6.8%) | 4,427  (6.8%) | 0.00 |
|  | Diagnosis of Erectile Dysfunction | 7,248  (9.0%) | 11,268  (8.9%) | 0.00 | 5,914  (9.0%) | 6,032  (9.2%) | 0.00 |

Table S3: Description of the tamsulosin versus 5ARI cohort before and after matching using propensity scores. Values reported are the number and percent (for categorical measures) and the median, lower, and upper quartiles (for continuous measures). The Std. Diff. column reports absolute value of standardized measures of differences between the 5ARI and tamsulosin groups. For continuous measures, the standardized difference is described using Cohen’s d. For categorical measures, Cohen’s w is used. Both Cohen’s d and w have roughly the same interpretation with values closer to 0 being smaller differences. By convention, standardized differences smaller than 0.1 are considered small-to-no difference. Before matching, tamsulosin users had later start dates, more lookback time, younger age and more less likely to have a diagnosis of abnormal PSA than users of 5ARI. After matching of the estimated standardized differences were below 0.1 with many being 0.01 or smaller.

|  |  | Entire Cohort | | | Propensity Score Matched Cohort | | |
| --- | --- | --- | --- | --- | --- | --- | --- |
|  |  | 5ARI | Tamsulosin | Std. Diff. | 5ARI | Tamsulosin | Std. Diff. |
| Sample Size, Number of Cases | Number of Enrollees | 80,158 | 437,045 | --- | 79,798 | 78,798 | --- |
|  | Number of Cases of DLB | 193 | 1,286 | --- | 192 | 224 | --- |
| Age, Timing of Index Date, Outcome | Age at Index Date | 62  (54-71) | 62  (56-71) | 0.10 | 62  (54-71) | 62  (54-71) | 0.00 |
|  | Index Date Year | 2010  (2008-13) | 2012  (2008-14) | 0.27 | 2010  (2008-13) | 2010  (2008-13) | 0.01 |
| Duration of Enrollment Time | Lookback Time (Years) | 3.2  (1.9-5.6) | 3.6  (2.0-6.3) | 0.18 | 3.2  (1.8-5.6) | 3.3  (1.9-5.8) | 0.06 |
|  | Follow-Up Time (Years) | 2.2  (1.0-4.4) | 1.9  (0.9-3.8) | 0.14 | 2.2  (1.0-4.4) | 2.2  (1.0-4.4) | 0.00 |
| Baseline Utilization and Complexity | Rate of Inpatient Events Per Year | 0.00  (0.00-0.00) | 0.00  (0.00-0.22) | 0.18 | 0.00  (0.00-0.00) | 0.00  (0.00-0.00) | 0.02 |
|  | Rate of Outpatient Events Per Year | 9  (5-16) | 10  (5-18) | 0.11 | 9  (5-16) | 9  (5-16) | 0.01 |
|  | Mean Number of Diagnosis Codes Per Outpatient Event | 1.40  (1.20-1.87) | 1.56  (1.27-2.09) | 0.24 | 1.40  (1.20-1.87) | 1.41  (1.20-1.88) | 0.01 |
| Elixhauser Comorbidities on Index Date | Alcohol Abuse | 1,152  (1.4%) | 11,379  (2.6%) | 0.03 | 1,152  (1.4%) | 1,221  (1.5%) | 0.00 |
|  | Anemia | 9,308  (11.6%) | 68,691  (15.7%) | 0.04 | 9,286  (11.6%) | 9,556  (12.0%) | 0.01 |
|  | Blood Loss | 913  (1.1%) | 7,232  (1.7%) | 0.01 | 913  (1.1%) | 956  (1.2%) | 0.00 |
|  | Heart Failure | 5,398  (6.7%) | 42,970  (9.8%) | 0.04 | 5,385  (6.7%) | 5,673  (7.1%) | 0.01 |
|  | Coagulopathy | 2,360  (2.9%) | 18,949  (4.3%) | 0.03 | 2,354  (2.9%) | 2,422  (3.0%) | 0.00 |
|  | Depression | 5,527  (6.9%) | 43.797  (10.0%) | 0.04 | 5,515  (6.9%) | 5,576  (7.0%) | 0.00 |
|  | Diabetes Without Complications | 15,970  (19.9%) | 121,292  (27.8%) | 0.06 | 15,959  (20.0%) | 16,249  (20.4%) | 0.00 |
|  | Diabetes with Complications | 5,141  (6.4%) | 46,695  (10.7%) | 0.05 | 5,137  (6.4%) | 5,247  (6.6%) | 0.00 |
|  | Drug Abuse | 576  (0.7%) | 6,690  (1.5%) | 0.02 | 575  (0.7%) | 621  (0.8%) | 0.00 |
|  | Fluid or Electrolyte Disorders | 6,001  (7.5%) | 56,021  (12.8%) | 0.06 | 6,994  (7.5%) | 6,370  (8.0%) | 0.01 |
|  | HIV | 548  (0.7%) | 1,438  (0.3%) | 0.02 | 516  (0.6%) | 547  (0.7%) | 0.00 |
|  | HTN Without Complications | 42,463  (53.0%) | 271,591  (62.1%) | 0.07 | 42,365  (53.1%) | 42,305  (53.0%) | 0.00 |
|  | HTN With Complications | 7,995  (10.0%) | 57,049  (13.1%) | 0.03 | 7,974  (10.0%) | 8,181  (10.3%) | 0.00 |
|  | Hypothyroidism | 7,062  (8.8%) | 43,432  (9.9%) | 0.01 | 7,037  (8.8%) | 7,268  (9.1%) | 0.01 |
|  | Liver Disease | 2,476  (3.1%) | 24,205  (5.5%) | 0.04 | 2,476  (3.1%) | 2,590  (3.2%) | 0.00 |
|  | Lymphoma | 844  (1.1%) | 6,367  (1.5%) | 0.01 | 841  (1.1%) | 915  (1.1%) | 0.00 |
|  | Metastatic Cancer | 1,108  (1.4%) | 10,716  (2.5%) | 0.03 | 1,105  (1.4%) | 1,184  (1.5%) | 0.00 |
|  | Neurological Disorders | 5,252  (6.6%) | 45,438  (10.4%) | 0.05 | 5,248  (6.6%) | 5,519  (6.9%) | 0.01 |
|  | Obesity | 4,530  (5.7%) | 48,019  (11.0%) | 0.06 | 4,529  (5.7%) | 4,649  (5.8%) | 0.00 |
|  | Paralysis | 1,006  (1.3%) | 10,583  (2.4%) | 0.03 | 1,005  (1.3%) | 1,077  (1.3%) | 0.00 |
|  | Pulmonary Hypertension | 1,592  (2.0%) | 13,310  (3.0%) | 0.02 | 1,591  (2.0%) | 1,713  (2.1%) | 0.01 |
|  | Psychoses | 4,257  (5.3%) | 32,490  (7.4%) | 0.03 | 4,240  (5.3%) | 4,355  (5.5%) | 0.00 |
|  | Peptic Ulcer Disease | 185  (0.2%) | 1,709  (0.4%) | 0.01 | 185  (0.2%) | 196  (0.2%) | 0.00 |
|  | COPD | 8,516  (10.6%) | 104,627  (23.9%) | 0.05 | 14,249  (17.9%) | 14,537  (18.2%) | 0.00 |
|  | Peripheral Vascular Disease | 3,543  (4.4%) | 65,114  (14.9%) | 0.04 | 8,504  (10.7%) | 8,773  (11.0%) | 0.01 |
|  | Renal Disease | 2,857  (3.6%) | 32,280  (7.2%) | 0.04 | 3,540  (4.4%) | 3,732  (4.7%) | 0.01 |
|  | Rheumatoid Arthritis | 2,857  (3.6%) | 22,205  (5.1%) | 0.03 | 2,854  (3.6%) | 2,882  (3.6%) | 0.00 |
|  | Solid Tumor | 10,567  (13.2%) | 73,745  (16.9%) | 0.04 | 10,564  (13.2%) | 11,084  (13.9%) | 0.01 |
|  | Valvular Disease | 9,339  (11.7%) | 62,332  (14.3%) | 0.03 | 9,317  (11.7%) | 9,525  (11.9%) | 0.00 |
|  | Weight Loss | 2,322  (2.9%) | 20,140  (4.6%) | 0.03 | 2,318  (2.9%) | 2,421  (3.0%) | 0.00 |
| Other Comorbidities and Treatment Selection Factors | Orthostatic Hypotension | 823  (1.0%) | 5,423  (1.2%) | 0.01 | 820  (1.0%) | 849  (1.1%) | 0.00 |
|  | Other Hypotension | 1,532  (1.9%) | 13,208  (3.0%) | 0.02 | 1,531  (1.9%) | 1,619  (2.0%) | 0.00 |
|  | PSA Measurement Taken | 48,463  (60.5%) | 249,926  (57.2%) | 0.02 | 48,194  (60.4%) | 48,184  (60.4%) | 0.00 |
|  | Diagnosis of Abnormal PSA | 23,789  (29.7%) | 70,538  (16.1%) | 0.13 | 23,516  (29.5%) | 23,465  (29.4%) | 0.00 |
|  | Diagnosis of Slow Urinary Stream | 1,963  (2.4%) | 13,752  (3.1%) | 0.01 | 1,960  (2.5%) | 2,019  (2.5%) | 0.00 |
|  | Uroflow Study Performed | 5,672  (7.1%) | 28,542  (6.5%) | 0.01 | 5,638  (7.1%) | 5,783  (7.2%) | 0.00 |
|  | Cystometrogram Performed | 546  (0.7%) | 3,485  (0.8%) | 0.00 | 546  (0.7%) | 543  (0.7%) | 0.00 |
|  | Diagnosis of BPH | 36,166  (45.1%) | 183,019  (41.9%) | 0.02 | 35,947  (45.0%) | 36,222  (45.4%) | 0.00 |
|  | Diagnosis of Anxiety | 5,501  (6.9%) | 39,365  (9.0%) | 0.03 | 5,484  (6.9%) | 5,519  (6.9%) | 0.00 |
|  | Diagnosis of Erectile Dysfunction | 7,248  (9.0%) | 45,596  (10.4%) | 0.02 | 7,219  (9.0%) | 7,288  (9.1%) | 0.00 |

S4: Coefficients of Cox regression with time-interacted covariates. Values in parenthesis are 95% CIs for the reported hazard ratio.

| Time | Tz/Dz/Az versus Tamsulosin | Tz/Dz/Az versus 5ARI | Tamsulosin versus 5ARI |
| --- | --- | --- | --- |
| 0-1 Years | 0.49 (0.33, 0.73) | 0.45 (0.26, 0.78) | 1.12 (0.77, 1.64) |
| 1-2 Years | 0.52 (0.33, 0.81) | 0.88 (0.50, 1.56) | 1.12 (0.70, 1.81) |
| 2-3 Years | 0.70 (0.43, 1.16) | 0.54 (0.28, 1.07) | 1.04 (0.62, 1.74) |
| 3-4 Years | 0.83 (0.48, 1.42) | 0.61 (0.31, 1.18) | 0.79 (0.45, 1.37) |
| 4-5 Years | 0.77 (0.44, 1.32) | 1.07 (0.53, 2.16) | 1.00 (0.57, 1.76) |
| 5-7.5 Years | 0.58 (0.37, 0.92) | 0.93 (0.45, 1.93) | 1.20 (0.66, 2.17) |
| 7.5-10 Years | 0.56 (0.32, 0.98) | 2.20 (0.76, 6.33) | 3.83 (1.56, 9.41) |
| > 10 Years | 0.50 (0.17, 1.46) | 0.66 (0.11, 3.97) | 3.01 (0.81, 11.2) |

S5: Cox proportional hazards estimates using BPH/LUTS sub-sample.

| Comparison | Sample Size | Hazard Ratio | 95% CI | p-value |
| --- | --- | --- | --- | --- |
| Tz/Dz/Az versus Tamsulosin | 89,188 | 0.73 | 0.55, 0.97 | 0.029 |
| Tz/Dz/Az versus 5ARI | 57,396 | 0.87 | 0.63, 1.20 | 0.409 |
| Tamsulosin versus 5ARI | 74,234 | 1.23 | 0.95, 1.59 | 0.117 |
